# Social contact patterns and their impact on the transmission of respiratory pathogens in rural China

**DOI:** 10.1101/2024.10.19.24315799

**Authors:** Yuxia Liang, Juanjuan Zhang, Qian You, Qianli Wang, Xiaohong Yang, Guangjie Zhong, Kaige Dong, Zeyao Zhao, Nuolan Liu, Xuemei Yan, Wanying Lu, Cheng Peng, Jiaxin Zhou, Jiqun Lin, Maria Litvinova, Mark Jit, Marco Ajelli, Hongjie Yu

## Abstract

**Introduction:** Social contact patterns significantly influence the transmission dynamics of respiratory pathogens. Previous surveys have quantified human social contact patterns, yielding heterogeneous results across different locations. However, significant gaps remain in understanding social contact patterns in rural areas of China.

**Methods:** We conducted a pioneering study to quantify social contact patterns in Anhua County, Hunan Province, China, from June to October 2021, when there were minimal coronavirus disease-related restrictions in the area. Additionally, we simulated the epidemics under different assumptions regarding the relative transmission risks of various contact types (e.g., indoor versus outdoor, and physical versus non-physical).

**Results:** Participants reported an average of 12.0 contacts per day (95% confidence interval: 11.3–12.6), with a significantly higher number of indoor contacts compared to outdoor contacts. The number of contacts was associated with various socio-demographic characteristics, including age, education level, income, household size, and travel patterns. Contact patterns were assortative by age and varied based on the type of contact (e.g., physical versus non-physical). The reproduction number, daily incidence, and infection attack rate of simulated epidemics were remarkably stable.

**Discussion:** We found many intergenerational households and contacts that pose challenges in preventing and controlling infections among the elderly in rural China. Our study also underscores the importance of integrating various types of contact pattern data into epidemiological models and provides guidance to public health authorities and other major stakeholders in preparing and responding to infectious disease threats in rural China.

## Introduction

Respiratory pathogens mainly spread through respiratory droplets when infected individuals cough, sneeze, speak, or breathe(1). The type of interaction between individuals, where the interaction takes place, the distance between individuals, and other factors play a role in shaping the transmission risk. For example, severe acute respiratory syndrome coronavirus 2 and Mycobacterium tuberculosis can spread directly with aerosols or droplets shed by an infector, specifically in poorly ventilated and/or crowded indoor settings(2). Therefore, most analyses of the transmission dynamics of respiratory pathogens require quantitative estimates of human social contacts and their characteristics. Many surveys have aimed to quantify human social contact patterns. The results were qualitatively similar between studies but depicted major quantitative differences(3-5), including between urban and rural areas(6, 7).

China has 1.4 billion residents in a vast area of about 9.6 million km^2^, with uneven economic development levels and varied modes of production and lifestyle. Such a heterogeneous population also translates into heterogeneous contact patterns, as exhibited in previous social contact surveys(8-14). Numerous contact surveys have been conducted in other parts of the world. However, there remain significant gaps in the knowledge of human social contact patterns in rural China. The only published social contact survey conducted in rural areas of Southern China was conducted in Guangzhou City (a city of over 18 million people and the capital of Guangdong province) in 2009–2010(15). To address this gap, we conducted a cross-sectional survey-based study in Anhua County, Hunan Province – a rural area in Central China – and explored the main determinants of contact patterns. Additionally, we conducted mathematical modeling analysis to assess the effect of contact patterns on the transmission dynamics of respiratory pathogens.

## Methods

### Study site and time

We conducted a contact survey from June to October 2021 in Anhua County, Yiyang City, Hunan Province, China. This county, located in Central China, is a highly rural area characterized by low income ($1164/year per capita for rural residents in 2021(16)) and low population density (158 people/km^2^ according to the 7^th^ China Population Census(17)). Since this study aimed to quantify social contact patterns under “normal” conditions, the survey was conducted when there were no widespread coronavirus disease (COVID-19) outbreaks or strict epidemic prevention and control measures. The Oxford COVID-19 Government Response Tracker (OxCGRT) index for Mainland China ranged from 67.13 to 79.17 during this time, which was higher than most countries in Europe and North America(18). During this period, Mainland China was classified into low-, medium-, and high-risk zones based on the COVID-19 situation. For low-risk areas, routine prevention measures were recommended, such as reducing gatherings and wearing masks indoors, but were not strictly enforced. In contrast, medium- and high-risk areas faced stricter measures, such as isolation and venue lockdowns(19). During the survey period, the entire Hunan Province was classified as a low-risk area(20).

### Study participants

We determined the minimum sample size based on the variance in the number of contacts per person per day, assuming a response rate of 90%. The minimum sample size was 828 and was allocated into eight age groups (0–9, 10–19, 20–29, 30–39, 40–49, 50–59, 60–69, and ≥ 70 years) according to the age structure of the Anhua population. Moreover, we doubled the sample size for individuals aged 0–19 to enhance parameter estimation accuracy, considering their significant role as the primary drivers of transmission for several infectious diseases(3). This resulted in a target sample size of 1,012 participants. Eligible participants included individuals of all ages who had resided in Anhua for more than three months in the year before the interview.

A probability proportional-to-population size sampling method was first applied, with three townships (Qingtang, Jiangnan, and Tianzhuang) in Anhua County, which were randomly selected for the study. In these three townships, we randomly selected 10 villages, 2 primary schools, and 3 middle schools, along with a high school in Anhua County. The residents of the 10 selected villages were approached in advance by local collaborators to obtain their consent to participate in the survey, resulting in a list of candidate households. Then, our interviewers visited those households one by one during the formal investigation. We recruited students by selecting particular classes based on the availability of students in those classes during the survey dates. Approximately 1–3 classes per school and 50 households per village were selected based on convenience sampling to broadly represent the entire Anhua population regarding geographical distribution. All students in the selected classes and all household members except students in the selected households were invited to participate in our study until we met our predefined target sample size for each age group.

### Data collection

School-aged children were enrolled at their schools, while other participants were recruited through door-to-door visits within the community. Well-trained interviewers administered face-to-face interviews, and questionnaires were completed by the interviewers. Participants were asked to retrospectively answer questionnaires about their social contact behaviors on two specific days: the most recent workday and the most recent weekend (i.e., the day before the interview and another most recent day with a different day type. If the day before the interview was a workday for the participant, the other day was the last weekend or holiday. If the day before the interview was a weekend or holiday for the participant, the other day was the last workday). Questionnaires were answered by guardians on behalf of children under 6 years or those who could not answer independently. The remaining participants answered the questionnaires by themselves. The questionnaire contained three main sections: (i) general information on the study participant, (ii) social contact behaviors on the latest non-holiday workday, and (iii) social contact behaviors on the latest weekend or holiday.

The general information included participants’ demographics, household composition, and travel behaviors.

Regarding social contact behaviors, contact is defined as either a two-way conversation involving three or more words in the physical presence of another person or direct physical contact (e.g., a handshake or hug)(3). We divided social contact into three categories: (i) physical contact, (ii) non-physical contact at a distance of 1 m or less, and (iii) non-physical contact at a distance of more than 1 m. We also recorded the age of the contactees, the relationship between the participant and contactee, and face mask usage during the contact (i.e., both participant and contactee wore a mask, only the participant, only the contactee, or none of them), contact location (i.e., indoor, outdoor, or both), contact setting (i.e., house, workplace, school, or community), distance between home and the furthest contact setting (i.e., < 1 km, 1–3 km, 4–5 km, 6–10 km, or > 10 km), and the frequency of contact (i.e., almost daily, 1–2 times per week, 1–2 times per month, less than once a month, or never met before).

Large numbers of contacts are difficult to record individually, which can lead to underreporting(14). This is similar to the results of previous studies(8, 9, 14, 21). Contacts with individuals sharing the same characteristics (e.g., a group of classmates) were recorded as a single entry, and the number of individuals in the group was also reported. We referred to this as group contact. Contacts with individuals with different characteristics were reported individually, and we referred to this as individual contact.

## Statistical analysis

### Covariates

Four types of explanatory variables were considered when describing human social contact. (i) Demographic and socioeconomic characteristics of the participants; specifically, sex, age, education level, occupation, individual income (in Chinese Yuan, CNY), household size, and number of years living in Anhua County (i.e., < 1 year, 1–5 years, 6–10 years, and > 10 years). (ii) Factors associated with the day the recorded contacts took place; specifically, type of the day (i.e., workday or weekend), whether the day is considered as typical by the participant (i.e., contact behavior similar to other workdays or weekends), and weather (i.e., sunny, cloudy, rainy, or varied). (iii) Whether participants regularly traveled out of the village where they reside daily or only occasionally. (iv) Other information about the participant; specifically, whether the participant has an underlying disease, contacts with animals (including raising, touching, or slaughtering animals), health status, and a self-rated contact memory accuracy (i.e., very well, well, moderate well, not well, or poorly).

### Characterization of human-to-human contact patterns

We used weighted generalized additive mixed models to explore the associations between participants’ age and the number of overall contacts per participant while controlling for the covariates mentioned above. We fitted penalized splines to explore the potential nonlinear relationships of continuous participants’ age to the response variable, considering the interactions between age and day type. The sampling structure of our study allowed us to include the random effect in our models to consider the proportion of variance in response variables attributable to intra- and inter-household and person variation. The regression model was weighted by the population size of each age group in Anhua County and defined as follows:

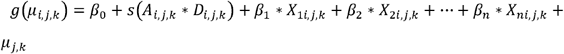

where *g* is the link function, and the response variable was assumed with a negative binomial distribution; *μ*_*i,j,k*_ is the number of overall contacts of *i*th contact diary reported by participant *j* from household k; *A*_*i,j,k*_ is the age of each participant; *D*_*i,j,k*_ is the day type of diary day; s(*A*_*i,j,k*_ * *D*_*i,j,k*_)denotes the nonlinear relationship between participants’ age and the response variable and the interactions between age and day type. We included this interaction because we found differences in the age distribution of contact between weekdays and weekends, specifically among those under 30 years (Figure S4). *X*_*1*_,…,*X*_*n*_ are other factors related to the number of overall contacts; *β*_1_,…,*β*_*n*_ are the partial regression coefficients of the above factors. *β*_0_ is the fixed intercept; *μ*_*j,k*_ is the intercept; *μ*_*i,j,k*_ = *E* (*Y*_*i,j,k*_ | *μ*_*j,k*_) is the expected value of *Y*_*i*_ under a given intercept. Besides exploring the determinants of contact patterns, we used the same model structure to test the effect of recall bias by comparing the number of contacts when respondents were asked to recall the day before the interview versus the most recent workday/weekend (i.e. an earlier day with a different day type), while accounting for various covariates.

To select the explanatory variables for the final model, we first fitted univariate regression models (generalized linear models with the same link functions as the multivariate model mentioned above) to identify candidate variables, using a significance level of 0.1. Next, we calculated Spearman’s correlation coefficient for each pair of candidate variables and excluded variables with a correlation threshold above 0.6. Finally, we used the multivariate regression model defined above, employed a bidirectional stepwise selection process based on Akaike’s Information Criterion to determine the final set of variables, and used a significance level of 0.05 in the multivariate regression models. We followed the same procedure to explore factors associated with the number of indoor and outdoor contacts.

### Contact matrices by age

We considered 17 age groups (0–2, 3–6, 7–9, 10–14, 15–19, …, and 75 years and above) to define age-specific contact matrices (*M*_*i,j*_) representing the average number of contacts per day that an individual in age group i have with individuals in age group j using the “socialmixr” package in R 4.3.2. We used a bivariate smoothing approach to estimate the elements of the contact matrices(3). The basis was a tensor-product spline ensuring flexibility when modeling the average number of contacts as a function of the participant’s and contact’s age over the 1-year band. The average number of contacts between a participant of age i and individuals of age j was modeled using a two-dimensional continuous function applied to the age of participants and contacts via a generalized additive model (GAM). To allow for over-dispersion in the number of contacts, we assumed that they were distributed following a negative binomial distribution with the mean *M*_*i,j*_, dispersion parameter *K*, and variance 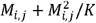. We used the basis dimension K = 10 to be large enough to fit the data well but small enough to keep the fitting procedure efficient. We then selected a thin plate regression spline to avoid the selection of the number of knots and a log link function for the GAM. Finally, we accounted for the reciprocal nature of the data and predicted the expected number of contacts at the population level.

### Transmission model

To estimate the impact of contact patterns on the transmission of respiratory pathogens, we used a deterministic age-structured compartmental Susceptible-Infected-Removed (SIR) model. Susceptible individuals were exposed to an age-specific force of infection regulated by the average number of contacts per day that an individual of a given age group had individuals of all age groups (i.e., the contact matrix estimated using the collected survey data). The model is described by the following set of differential equations:

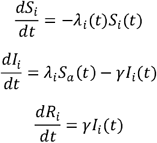

where *i* represents the age group (0–2, 3–6, 7–9, 10–14, 15–19, …, and 75 years and above); *S*_*i*_ (*t*) is the number of susceptible individuals in age group *i* at time t; *I*_*i*_ (*t*) is the number of infected individuals in age group *i* at time *t*; *R*_*i*_ (*t*) is the number of removed individuals in age group *i* at time t, and *γ* represents the removal rate, which corresponds to the inverse of the generation time in a SIR model(22).

The force of infection to which age group i is exposed to, denoted with *λ*_*i*_ (*t*), is defined as follows:

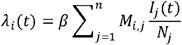

where *N*_*i*_ is the number of individuals in age group *i*, and *β* represents the per-contact transmission risk, and it is determined to obtain the desired value of the basic reproduction number (*R*_0_) using the next-generation matrix (NGM) approach: 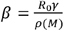 where *ρ* (*M*)represents the spectral radius of contact matrix *M*.

In this study, we did not aim to simulate a specific respiratory pathogen or model a particular epidemic phase. Therefore, as an illustrative scenario, we assumed *R*_0_ = 2 and the generation time to be 5.1 days, both in line with values observed during the COVID-19 pandemic(23, 24). Moreover, to represent the transmission of pathogens with potentially different characteristics, we investigated scenarios where we altered: (i) the transmission risk of outdoor relative to indoor contacts (namely, 10%, 50%, and 90%); (ii) non-physical relative to physical contacts (namely, 10%, 50%, and 90%); and (iii) non-physical contacts at more than 1-meter distance relative to physical or non-physical contacts at less than 1-meter distance (namely, 10%, 50%, and 90%). Then, we estimated the reproduction number based on the relative transmission risk of different types of contacts. The number of individuals by age corresponded to that of the population in Anhua County (780,969 individuals). Model simulations were initialized with one infected individual per hundred thousand, and all other individuals were considered susceptible. The model was run for 180 days, and epidemic sizes obtained using different contact matrices were compared.

## Ethical approval

All procedures performed in this study were approved by the ethics committees of the School of Public Health at Fudan University, Shanghai, China (Ref: 2020-11-0857-S2) and the London School of Hygiene and Tropical Medicine, London, UK (Ref: 22684-04), and were following the 1964 Helsinki declaration and its later amendments or comparable ethical standards. Informed consent was obtained from all individual participants included in the study (from a guardian if the participant was under 6 years old; from both a guardian and the study participant if the participant was 7–17 years old; from the participant if the participant was 18 years or above).

## Results

### Analyzed sample

A total of 1,522 participants were recruited for this study. The data analysis was based on a sample of 1,499 participants, and 23 (1.5%) were excluded due to data quality issues (Figure S1). Among the participants, 575 (38.4%) were male, and the average age was 29. A comparison between the characteristics of the study participants and those of the Anhua County population is reported in Table S1 and Figure S2. For each participant, we recorded their contact behaviors for two days.

Overall, 2912 contact diaries (1496 from the day before the interview and 1416 from the other day) (Figure S3) were collected, corresponding to 34,802 contacts, which consisted of 33,609 recorded contacts with details and 1193 contacts that respondents estimated they had left out. When the survey was conducted, there were no statutory holidays and no reports of COVID-19 cases in Anhua County and Yiyang City, with 3 local COVID-19 cases in Hunan Province and fewer than 15 new COVID-19 cases daily in Mainland China. No stringent epidemic prevention and control measures were in place(25). From June to July 2021, the daily maximum temperatures in Anhua County ranged between 21 and 36 □; in October 2021, the daily maximum temperatures ranged from 11 to 21 □ (Figure S3)(26).

### Number of contacts

According to the results of the recall bias test, the number of contacts on the other day was 1.1 times that of the day before the interview after correcting for covariates, with an absolute mean increase of 0.9 contacts per person per day (Table S2), suggesting that the recall bias did not have a large effect on the results. The frequency distribution of the number of contacts was right-skewed (Figure 1A). The mean number of contacts was 12.0 (95% confidence interval [CI]: 11.3–12.6), the median number of contacts was 7 (interquartile range: 4–14), and the number of contacts was heterogeneous according to participant characteristics (Tabs. 1 and S3 and Figure 1B). For example, participants aged 7–19 reported the highest number of contacts with 15.4 (95% CI: 14.4–16.4). The mean number of contacts for individuals under 30 years exhibited notable differences between weekends and workdays, whereas for those over 30 years, the differences almost disappeared (Figure S4). Participant characteristics also influence the distribution of contacts by social setting. For instance, children aged 0–2 years had most of their contacts at home (69.0%), while participants aged 7–19 had most of their contacts at school (75.2%) (Figure S5A). Face-mask usage was low, with 93.7% of contacts occurring without masks (Figure S5C). Across all age groups, most contacts occurred within a 1 km radius of the participants’ homes and involved individuals they met almost daily (Figure S5D-E). Of the 34,802 contacts, 27,419 indoor, 6,078 outdoor, and 1,305 contacts with unknown settings were recorded. Indoor contacts were more prevalent across all age groups, with 9.4 (95% CI: 9.0–9.9) contacts per day as compared to 2.1 (95% CI: 1.8–2.4) outdoor contacts per day (Table 1). Indoor contacts mainly took place at school for participants aged 7–19 (76.0%) and at the workplace for those aged 20–39 (44.8%). For the other age groups, the home was the primary setting for indoor contact (Figure S5F). Outdoor contact primarily occurred in the community setting for most age groups (Figure S5K). Physical contact was reported in 38.2% of the interactions, with the proportion of physical contact decreasing with participant age. Individuals aged 18 years and younger reported more physical contact than non-physical ones, whereas adults had a higher proportion of non-physical contacts (Figure 1C-D).

**Table 1.** Mean (95%CI) number of contact diaries and mean number of total, indoor, and outdoor contacts by participant’s characteristic (see Table S3 for median and IQR).

| Characteristics | N (%) <sup>a</sup> | Mean (95%CI) <sup>b</sup> |  |  |
| --- | --- | --- | --- | --- |
|  |  | Total | Indoor | Outdoor |
| **Overall** | 2912 (100) | 12.0 (11.3, 12.6) | 9.4 (9.0, 9.9) | 2.1 (1.8, 2.4) |
| **Sex** |  |  |  |  |
| Male | 1108 (38.0) | 11.7 (10.8, 12.8) | 8.9 (8.2, 9.6) | 2.4 (1.8, 3) |
| Female | 1804 (62.0) | 12.1 (11.4, 12.9) | 9.7 (9.2, 10.4) | 1.9 (1.6, 2.3) |
| **Age group** |  |  |  |  |
| 0-2 yrs | 214 (7.3) | 6.0 (5.5, 6.4) | 5 (4.7, 5.3) | 0.9 (0.6, 1.2) |
| 3-6 yrs | 343 (11.8) | 10.4 (9.2, 11.6) | 9 (7.9, 10.2) | 1.3 (0.9, 1.7) |
| 7-19 yrs | 660 (22.7) | 15.4 (14.4, 16.4) | 13.4 (12.5, 14.4) | 0.6 (0.5, 0.8) |
| 20-39 yrs | 685 (23.5) | 14.5 (12.8, 16.5) | 10.7 (9.6, 12) | 3.7 (2.6, 5.1) |
| 40-59 yrs | 690 (23.7) | 11.3 (10.3, 12.6) | 8 (7.2, 9.1) | 2.9 (2.4, 3.4) |
| 60-75 yrs | 254 (8.7) | 6.6 (5.8, 7.4) | 4.9 (4.3, 5.7) | 1.5 (1.2, 1.9) |
| ≥76 yrs | 66 (2.3) | 5.6 (4, 7.7) | 3.4 (2.9, 4) | 2 (0.5, 3.7) |
| **Education** |  |  |  |  |
| None | 1133 (38.9) | 8.9 (8.3, 9.5) | 7.3 (6.8, 7.8) | 1.5 (1.3, 1.8) |
| Primary school | 670 (23.0) | 11.5 (10.4, 12.7) | 8.4 (7.4, 9.4) | 2.1 (1.7, 2.4) |
| Middle school | 711 (24.4) | 13.4 (12.5, 14.3) | 11.4 (10.5, 12.2) | 1.5 (1.1, 1.8) |
| High school | 232 (8.0) | 13.1 (10.7, 15.5) | 10.3 (8.4, 12.9) | 2.6 (1.5, 4) |
| College or above | 166 (5.7) | 26.6 (20.6, 32.8) | 18.4 (15, 21.8) | 8 (3.7, 13.6) |
| **Occupation** |  |  |  |  |
| Enrolled in pre-kindergarten | 310 (10.6) | 6.4 (5.9, 6.9) | 5.1 (4.9, 5.4) | 1.1 (0.8, 1.5) |
| Enrolled in kindergarten | 212 (7.3) | 12.2 (10.5, 13.9) | 11.1 (9.5, 12.8) | 0.9 (0.6, 1.4) |
| Student | 691 (23.7) | 14.8 (13.8, 15.8) | 12.9 (12, 13.7) | 0.7 (0.6, 0.9) |
| Self-employed | 86 (3.0) | 20.4 (16.9, 24.2) | 15.5 (12.2, 19.4) | 2.8 (1.4, 4.5) |
| Farmer | 754 (25.9) | 8.3 (7.5, 9.2) | 5.9 (5.2, 6.7) | 2.2 (1.9, 2.6) |
| School staff | 70 (2.4) | 33.4 (22.9, 46.9) | 25.7 (20, 32.1) | 7.6 (2, 16.6) |
| Other staff | 72 (2.5) | 29.9 (21.5, 40.8) | 16.3 (12.1, 21.2) | 12.9 (5, 22.2) |
| Others | 327 (11.2) | 14.2 (12.3, 16.2) | 11.4 (9.7, 13.4) | 2.5 (2, 3.1) |
| Unemployed | 390 (13.4) | 7.4 (6.8, 8.1) | 5.3 (4.9, 5.8) | 2.1 (1.6, 2.6) |
| **Individual income (CNY)** |  |  |  |  |
| No income | 342 (11.7) | 7.5 (6.8, 8.2) | 5.2 (4.9, 5.6) | 2.2 (1.7, 2.8) |
| < ¥1 k | 20 (0.7) | 7.0 (4.1, 11.5) | 5.9 (3, 10.2) | 1.1 (0.4, 2) |
| ¥1-9 k | 166 (5.7) | 10.3 (8.8, 12) | 6.7 (5.5, 8.1) | 3.2 (2.3, 4.3) |
| ¥10-29 k | 322 (11.1) | 14.3 (12.6, 16.3) | 10 (8.7, 11.5) | 3.6 (2.6, 4.8) |
| ≥ ¥30 k | 291 (10) | 20.8 (17.1, 24.9) | 15.6 (13.2, 18.2) | 5 (2.4, 8) |
| Not available | 1213 (41.7) | 12.2 (11.5, 12.8) | 10.6 (10, 11.2) | 0.9 (0.7, 1) |
| Unwilling to answer | 531 (18.2) | 8.6 (7.5, 9.8) | 6.6 (5.6, 7.7) | 2 (1.6, 2.4) |
| Unknown/no response | 27 (0.9) | 13.3 (7.5, 19.4) | 11.7 (6.3, 18.2) | 1.6 (0.5, 2.9) |
| **Household size** |  |  |  |  |
| 1-3 | 1216 (41.8) | 10.1 (9.2, 11) | 7.4 (6.7, 8.2) | 2.3 (1.8, 2.9) |
| 4-6 | 1258 (43.2) | 12.3 (11.5, 13.3) | 9.8 (9.1, 10.5) | 2.2 (1.7, 2.8) |
| ≥7 | 436 (15) | 16.2 (15.0, 17.3) | 14 (13, 15) | 1.2 (0.9, 1.6) |
| Unknown/no response | 2 (0.1) | 5.0 (5.0, 5.0) | 1 (1, 1) | 4 (4, 4) |
| **Number of years living in Anhua county** |  |  |  |  |
| <1 yrs | 56 (1.9) | 8.1 (6.4, 10.4) | 7.2 (5.6, 9.6) | 0.8 (0.3, 1.3) |
| 1-5 yrs | 507 (17.4) | 9.0 (8.3, 10.0) | 7.9 (7.1, 8.8) | 1 (0.7, 1.3) |
| 6-10 yrs | 327 (11.2) | 11.7 (10.6, 13.0) | 9.8 (8.8, 10.8) | 1.3 (0.8, 1.8) |
| >10 yrs | 2004 (68.8) | 12.8 (12.1, 13.7) | 9.8 (9.2, 10.5) | 2.5 (2.1, 3.1) |
| Unknown/no response | 18 (0.6) | 12.4 (9.0, 16.4) | 9.1 (5.9, 13.2) | 1.3 (0.6, 2.2) |
| **Day type** |  |  |  |  |
| Workday | 1424 (48.9) | 14.5 (13.5, 15.6) | 11.6 (10.8, 12.4) | 2.3 (1.8, 3) |
| Weekend | 1488 (51.1) | 9.5 (8.9, 10.2) | 7.3 (6.9, 7.8) | 1.9 (1.5, 2.3) |
| **Typical day** |  |  |  |  |
| Yes | 2656 (91.2) | 11.5 (10.8, 12.1) | 9 (8.6, 9.5) | 2 (1.7, 2.4) |
| No | 239 (8.2) | 17.5 (14.9, 20.2) | 13.9 (11.6, 16.6) | 3.1 (2.2, 4.2) |
| Unknown/no response | 17 (0.6) | 11.7 (6.5, 17.9) | 8.7 (4.2, 13.7) | 1.1 (0.3, 2) |
| **Weather** |  |  |  |  |
| Sunny | 1739 (59.7) | 11.7 (10.9, 12.6) | 8.7 (8.2, 9.3) | 2.5 (2, 3) |
| Cloudy | 361 (12.4) | 11.8 (10.3, 13.3) | 9.8 (8.7, 11.1) | 1.7 (1, 2.6) |
| Rainy | 663 (22.8) | 12.7 (11.8, 13.7) | 10.9 (9.9, 11.9) | 1.4 (1.1, 1.8) |
| Variable | 101 (3.5) | 13.2 (9.9, 17.9) | 11.5 (8.1, 15.5) | 1.5 (0.9, 2.1) |
| Unknown/no response | 48 (1.6) | 9.0 (5.6, 13.9) | 6.9 (4.4, 10.8) | 1.2 (0.5, 2) |

| Frequency of travels outside the village of residence |  |  |  |  |
| --- | --- | --- | --- | --- |
| Almost daily | 329 (11.3) | 19.2 (16.2, 22.9) | 14 (12.2, 16.2) | 4.4 (2.3, 7.1) |
| At least once a week but not daily | 586 (20.1) | 12.9 (11.7, 14.1) | 10.1 (9.1, 11.1) | 2.1 (1.6, 2.7) |
| At least once a month but not each week | 777 (26.7) | 11.8 (10.8, 12.9) | 9.8 (8.8, 11) | 1.6 (1.4, 2) |
| Less than monthly, but not never | 925 (31.8) | 9.7 (9.0, 10.4) | 7.5 (7, 8.1) | 1.9 (1.5, 2.3) |
| Never | 239 (8.2) | 7.7 (6.7, 8.7) | 6.2 (5.3, 7.3) | 1.2 (0.8, 1.8) |
| Unknown/no response | 56 (1.9) | 17.4 (14.3, 20.8) | 14.3 (11.4, 17.6) | 1.5 (0.7, 2.5) |
| Underlying conditions |  |  |  |  |
| Yes | 597 (20.5) | 10.5 (9.2, 12.1) | 7.3 (6.5, 8.3) | 3 (2.1, 4.1) |
| No | 2200 (75.5) | 12.3 (11.7, 13.1) | 10 (9.4, 10.5) | 1.9 (1.6, 2.3) |
| Unknown/no response | 46 (1.6) | 14.9 (11.0, 20.1) | 12.7 (9.1, 17.7) | 0.1 (0, 0.2) |
| Unwilling to answer | 69 (2.4) | 9.6 (7.5, 12.1) | 8 (6.3, 9.8) | 0.7 (0.3, 1.1) |
| Contact with animals |  |  |  |  |
| Yes | 1713 (58.8) | 10.9 (10.2, 11.7) | 8.4 (7.8, 9) | 2.3 (1.9, 2.8) |
| No | 1195 (41.0) | 13.4 (12.6, 14.4) | 11 (10.3, 11.7) * | 1.7 (1.3, 2.4) * |
| Unknown/no response | 4 (0.1) | 7.5 (5.0, 10.0) | 3.5 (1, 6) | 4 (4, 4) |
| Self-rated contact memory accuracy |  |  |  |  |
| Very well | 1439 (49.4) | 10.0 (9.3, 10.8) | 7.9 (7.4, 8.5) | 2.1 (1.7, 2.6) |
| Well | 1021 (35.1) | 12.2 (11.2, 13.4) | 9.8 (9.2, 10.6) | 1.9 (1.4, 2.6) |
| Moderate well | 390 (13.4) | 18.0 (16.1, 20.3) | 14.1 (12.4, 16.2) | 2.3 (1.5, 3.3) |
| Not well | 45 (1.5) | 16.8 (12.8, 20.9) | 7.4 (5.8, 9.1) | 5.4 (2.5, 8.8) |
| Unknown/no response | 17 (0.6) | 13.0 (6.6, 19.4) | 10.3 (4.8, 16.2) | 0.6 (0.1, 1.4) |
| Health status |  |  |  |  |
| Very well | 1973 (67.8) | 11.9 (11.2, 12.7) | 9.3 (8.8, 9.9) | 2.2 (1.8, 2.7) |
| Well | 641 (22.0) | 12.2 (11.1, 13.2) | 10 (9.1, 11) | 1.6 (1.3, 2) |
| Moderate well | 229 (7.9) | 11.8 (10.2, 13.5) | 9.1 (7.8, 10.5) | 2.2 (1.6, 2.9) |
| Not well | 51 (1.8) | 11.5 (7.7, 15.5) | 7.1 (4.6, 10.1) | 3.7 (1.4, 6.5) |
| Unknown/no response | 18 (0.6) | 11.3 (6.4, 16.7) | 8.4 (4.3, 13.1) | 0.7 (0.1, 1.4) |

<sup>a</sup> Number of contact diaries.
<sup>b</sup> Based on 100 bootstrap samples.

**Figure 1.**
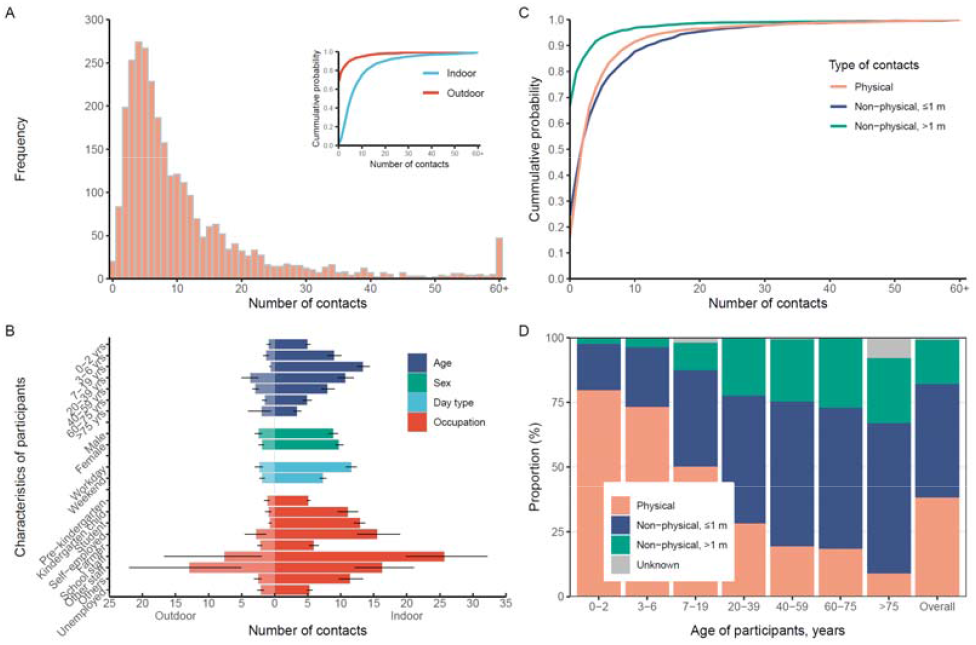
Distribution of the number of contacts. **(A)** Frequency of the number of contacts. The inset depicts the cumulative probability distribution disaggregated by indoor and outdoor contacts. **(B)** Mean and 95% CI of the number of indoor (right) and outdoor (left) contacts by participants’ characteristics. **(C)** Cumulative probability distribution of the number of contacts by type of contact (namely, physical, non-physical at ≤ 1 m distance, and non-physical at > 1 m distance). **(D)** The proportion of contacts by contact type disaggregated by study participant age.

### Determinants of contact patterns

Our statistical model illustrated different nonlinear patterns on workdays and weekends. Individuals under 30 years had more contacts on workdays than on weekends. However, for those over 30 years, the number of contacts was similar on workdays and weekends. We found a significant nonlinear association between the number of contacts and the age of participants, with the highest number of contacts for respondents aged approximately 20 years on workdays (Figure 2A). Workdays, higher level of education, higher individual income, larger household size, worse self-rated contact memory accuracy, and daily travel out of the village of residence were significantly associated with a larger number of contacts (Figure 2B).

**Figure 2.**
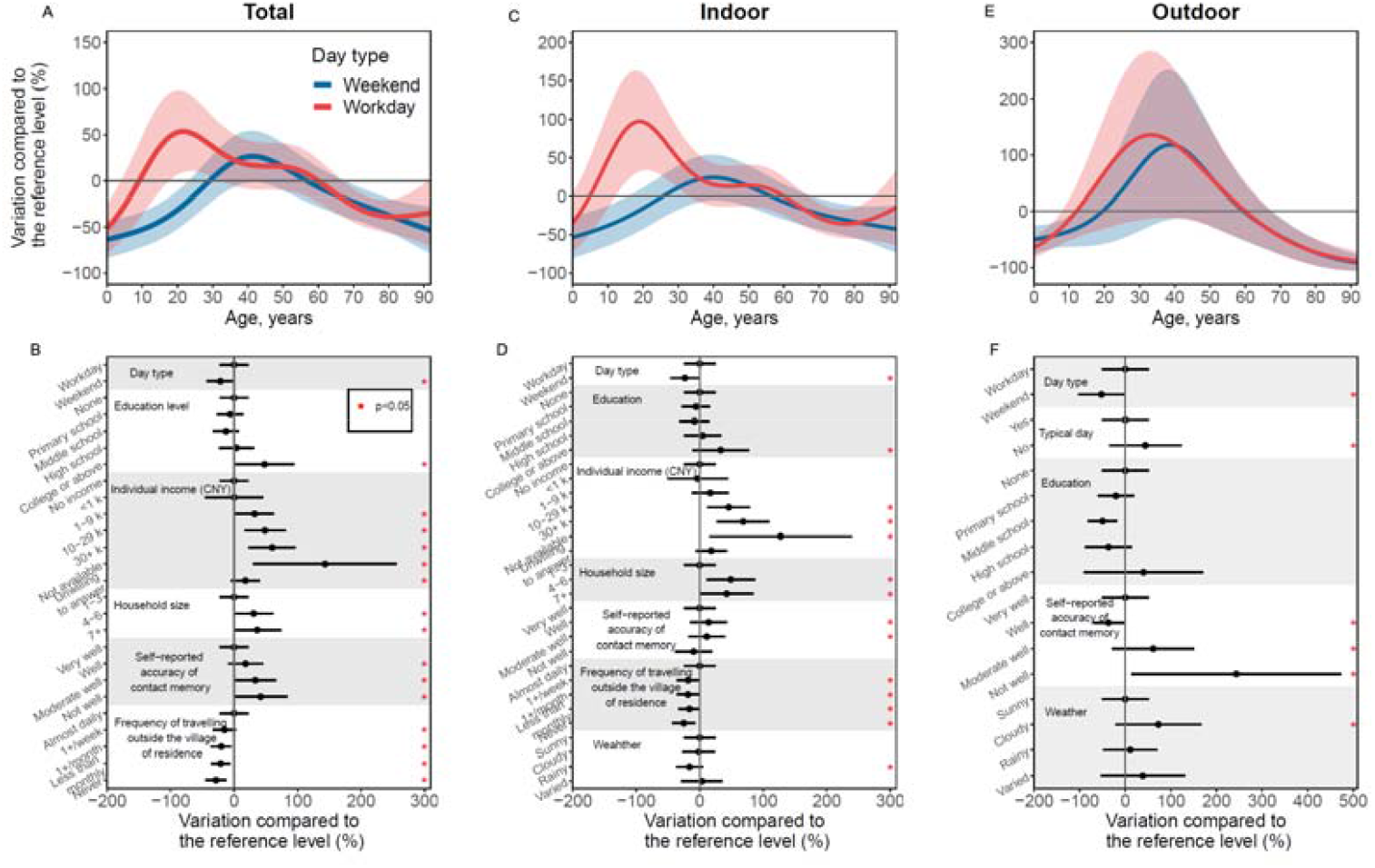
Determinants of contact patterns. **(A)** Variation by age in the number of total contacts compared to the reference level. **(B)** Variation by factors other than age in the number of total contacts compared to the reference level. **(C)** Same as **A**, but for the number of indoor contacts. **(D)** Same as **B**, but for the number of indoor contacts. **(E)** Same as **A**, but for the number of outdoor contacts. **(F)** Same as **B**, but for the number of outdoor contacts. Reference level means 60-year-old participants with other factors at the first level. For example, for panels **(A)** and **(B)**, the reference level means 60-year-old participants on workdays, with non-education, no income, household size is 1–3, self-reported accuracy of contact memory being very well, and traveling outside the village of residence almost daily.

The number of indoor contacts was the highest for respondents aged around 20 years on workdays and around 40 years on weekends (Figure 2C). Workdays, higher level of education, higher individual income, larger household size, worse self-rated contact memory accuracy, and daily travel out of the village of residence were significantly associated with larger number of indoor contacts. Moreover, rainy days were associated with fewer indoor contacts compared to sunny days (Figure 2D). The number of outdoor contacts was the highest for respondents aged around 30–50 years both on workdays and weekends (Figure 2E). Workdays, typical days, worse self-rated contact memory accuracy, and cloudy weather were significantly associated with larger number of outdoor contacts (Figure 2F).

### Contact patterns by age

A prominent feature of the contact matrix by age was the presence of a strong diagonal, indicating that participants in all age groups mixed assortatively by age, with more than 85.0% of all contacts occurring within the same age group. This assortativeness was most pronounced in the 15–19-year age group (Figure 3 and Figure S6). The bottom-left corner of the matrices, corresponding to contacts between school-age children, recorded the largest number of contacts. We also found intergenerational contacts, such as contacts between those aged 0–10 years and those aged 20–30, contacts between those aged 20–30 and those aged 50–60, or contacts between those aged 0–10 and those aged 50–60 (Figure 3A). For those aged 7–19, social contacts mainly occurred in schools, while adults aged 20 years and above primarily made contacts in workplaces or communities (Figs. S7A–D). The indoor contact matrix was similar to the overall contact matrix, albeit with relatively fewer contacts (Figure 3B). The outdoor contacts were assortative by age (Figure 3C) and mainly occurred in the community (Figure S7I–L). We found similar mixing patterns by age between weekends and weekdays for adults over 30 years. However, individuals under 30 had fewer contacts during the weekend, specifically with individuals in the same age group (Figure S8). This difference is mainly evident for outdoor contacts. Physical contacts mainly occurred between people of the same age and between those aged 0–10 and 20–60, primarily corresponding to the contact matrices in households and schools (Figure 3D). Non-physical contacts at less than 1-meter distance are predominantly found among people aged 7–25 years and among people aged 30 years and above, mainly occurring in schools, workplaces or communities (Figure 3E).

**Figure 3.**
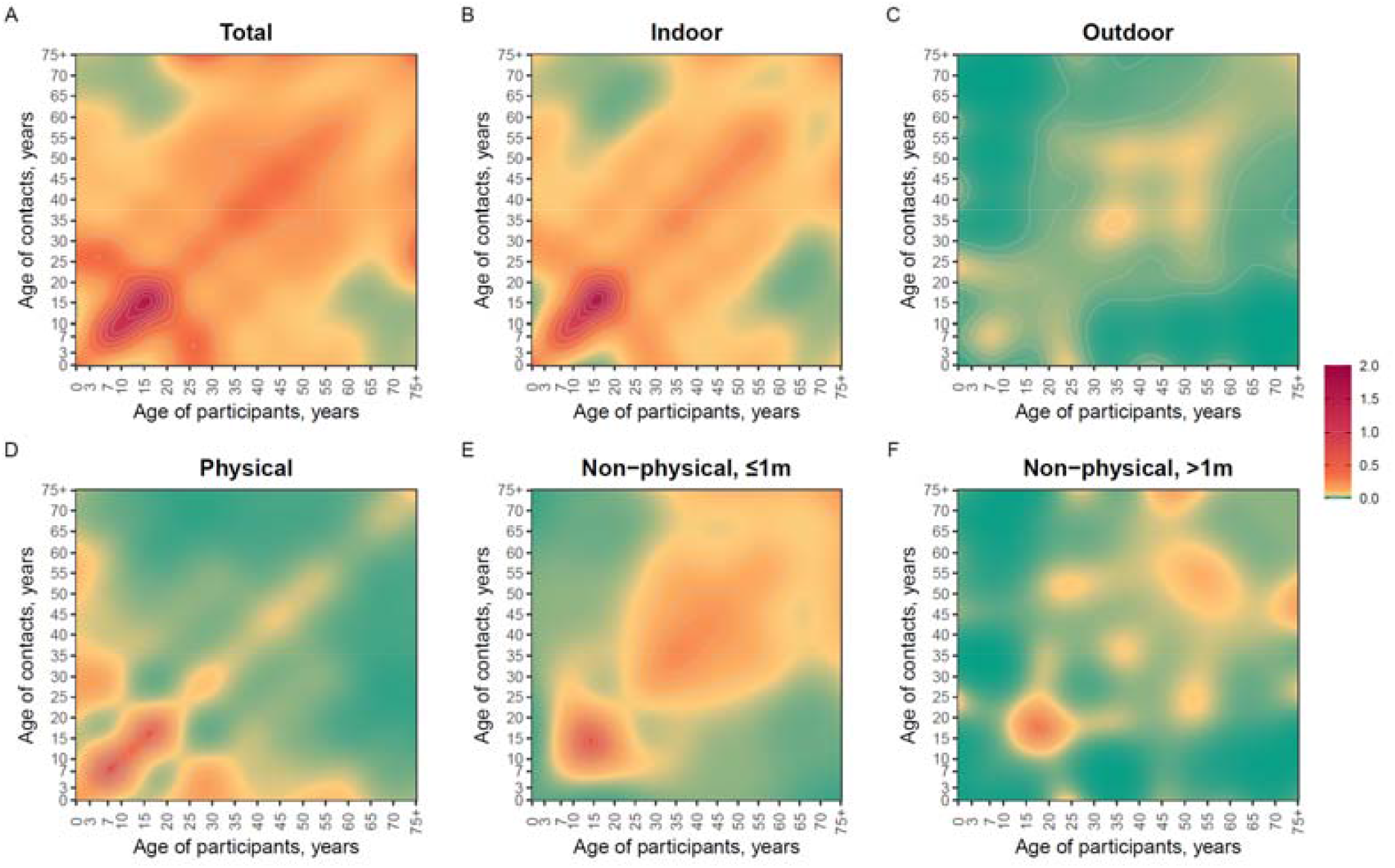
Smoothed symmetric contact matrices by age. **(A)** Total contact. **(B)** Indoor contact. **(C)** Outdoor contact. **(D)** Physical contact. **(E)** Non-physical contact at less than 1-meter distance **(F)** Non-physical contact at more than 1-meter distance.

### Contact patterns and transmission risk

In our baseline analysis, we assumed that all contacts had the same transmission risk. By setting *R*_0_ = 2, our mathematical model estimated a final infection attack rate of 54.5% (95% CI: 47.3%–60.7%) (Figure 4A-B). By considering a 10%–90% reduction in transmission risk for outdoor contacts relative to indoor ones, we estimated a 0%–5.0% reduction in the reproduction number, which led to an 11.7%–36.5% reduction in the final infection attack rate. Similar reductions in the estimated reproduction number and final infection attack rate were estimated when considering a 10%–90% reduction in transmission risk for non-physical contacts at more than 1-meter distance relative to physical or non-physical contact at less than 1-meter distance (Figure 4A-B). A more marked reduction was observed when considering a 10%–90% reduction in transmission risk for non-physical contacts relative to physical ones. Specifically, we estimated the reproduction number to decrease by 5.0%–45.0%, leading to a 13.6%–99.1% decrease in the final infection attack rate. When considering the relatively lower transmission risk for outdoor contacts, non-physical contacts, or non-physical contacts at more than 1-meter distance, lower peak incidences were observed (Figure 4C). Regarding the infection attack rate by age, all the analyzed scenarios led to the same qualitative trend with higher infection rates in younger individuals. However, quantitative differences exist depending on the assumed scenarios regarding the relative transmission risk by contact type (Figure 4D and S9).

**Figure 4.**
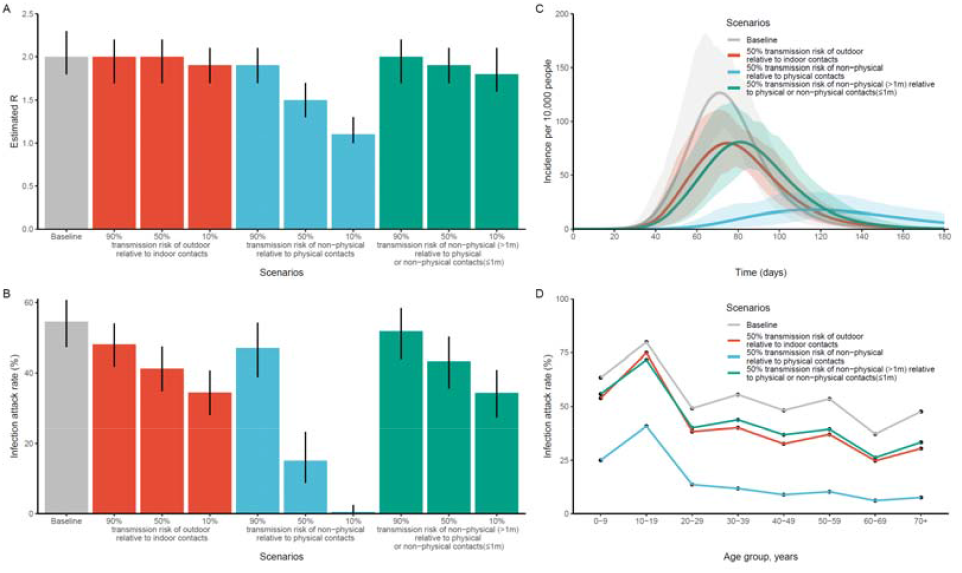
Effects of different assumptions on the relative transmission risk of different contact types. **(A)** Estimated *R* based on different scenarios of relative transmission risk of different types of contact. **(B)** Estimated infection attack rate based on different scenarios of relative transmission risk of different types of contact. **(C)** Estimated daily incidence based on different scenarios of relative transmission risk of different types of contact. **(D)** Estimated infection attack rate by age group based on different scenarios of relative transmission risk of different types of contact.

## Discussion

We conducted a social contact survey from June to October 2021 in Anhua County, Hunan Province, a rural area in Central China. Our findings depicted that participants reported an average of 12.0 (95% CI: 11.3–12.6) contacts per day, with significantly more indoor than outdoor contacts. The number of contacts was associated with several socio-demographic characteristics of the study participant, including age, level of education, income, household size, and travel patterns. Contact patterns were assortative by age and differed based on the type of contact (e.g., physical versus non-physical). Finally, we estimated the reproduction number, the infection attack rate, and the daily incidence of simulated stylized epidemics of generic respiratory pathogens to be remarkably stable under different assumptions on the relative transmission risk of various types of contacts (e.g., indoor versus outdoor, physical versus non-physical).

Our study is the first to quantify human-human contact patterns in rural areas of Central China. We compared our results to other contact studies conducted in rural areas and other studies in China in urban and rural areas. In comparison to surveys in rural areas, our mean daily contact (12.0) was similar to studies in Peru (12.0)(27), Zimbabwe (10.8)(6), and England (10.5)(28), which was higher than those in Ethiopia (5.73-6.19)(29), Vietnam (7.7)(30), Uganda (7.2)(31), and lower than those in rural Guangzhou (18.6)(15), Kenya (17.7)(32), and India (21.4)(33). Compared with other Chinese studies, we found fewer contacts than studies in Shanghai (18.9)(14), urban Guangzhou (18.6)(15), and Wuhan (14.6)(9), similar numbers to studies in urban Taiwan (12.5)(10), and higher numbers than those in Shenzhen (7.9)(8), Changsha (9.5)(8), and Hong Kong (8.1)(11). The differences could result from the combined effects of study location, study population, and survey methods. Additionally, we identified typical age assortativity in age-specific contact matrices, where individuals predominantly interact with others of a similar age. However, strong intergenerational contacts were observed within the population, consistent with findings from previous studies in rural areas(7, 33) and other regions of China(8, 9, 14, 21, 34, 35), but contrasting with the infrequency of such interactions in European contact matrices(3). This discrepancy may be attributed to cultural differences, as people in rural areas and China are more likely to have adults caring for older family members and grandparents caring for grandchildren(36). This may lead to older adults in rural areas having a higher risk of acquiring close-contact infections from younger people compared to those in other regions.

Contact patterns varied between workdays and weekends, primarily for students and a small portion of workers. However, for most participants, the patterns remained similar throughout the week. This consistency is likely due to the unique characteristics of agricultural and rural lives in Central China. In contrast, previous studies conducted in urban areas within and outside China found more pronounced differences between workdays and weekends(3, 14, 21). Despite the analysis being conducted in a rural area, we estimated a significantly higher number of indoor than outdoor contacts. This is particularly relevant for studying respiratory pathogens, as they spread predominantly indoors(37-40).

According to our mathematical modeling analysis, heterogeneity in contact patterns across different age groups, social settings, and environments translates into heterogeneous epidemiological outcomes. Our analysis also exhibits the importance of obtaining reliable estimates of the relative transmission risk of different types of contacts (e.g., physical versus non-physical contacts or indoor versus outdoor contacts) and the effectiveness of various interventions in reducing different types of contacts. Indeed, considering such differences could have a remarkable impact in estimating the effectiveness of public health interventions targeting certain social settings (e.g., museums and restaurants) or restricting certain behaviors (e.g., being at least 1 meter apart).

To interpret our findings properly, it is crucial to consider the limitations of our analysis. First, asking participants to report their contact behaviors retrospectively can introduce recall bias. The results of the multivariate analysis support this possibility, as we found that self-rated contact memory accuracy was negatively associated with the number of contacts. Likely, participants with numerous contacts struggled to recall all individuals they encountered, leading to lower self-ratings of their contact memory. Second, we defined group contacts to record interactions with individuals sharing identical characteristics systematically. While this approach may have reduced the burden on the study participants, it likely came at the cost of reduced accuracy in the information collected for each contact. The participants may have grouped contacts with similar rather than identical characteristics. Third, mathematical modeling analysis represents a simplified illustrative example of the spread of respiratory pathogens with potentially different characteristics. As such, it does not consider other nuances that may shape the transmission process, such as contact duration, susceptibility to infection by age, and waning immunity.

In conclusion, our analysis provides new knowledge on contact patterns in rural areas of Central China that are relevant to the transmission of respiratory pathogens. Future integration of contact pattern data with local epidemiological surveillance and seroepidemiological data could represent the next step to improve our understanding of the spread of respiratory pathogens. This could also provide major public health benefits by creating the capacity to prevent better, prepare, and respond to infectious disease threats.

## Data Availability

All individual-level data will be made available upon acceptance of the manuscript.

## Conflict of interests

HY has received research funding from Sanofi Pasteur, GlaxoSmithKline, Shanghai Roche Pharmaceutical Company, Shenzhen Sanofi Pasteur Biological Products Co., Ltd and SINOVAC Biotech Ltd.

## Funding

This study was funded by the Shanghai Municipal Science and Technology Major Project (ZD2021CY001), NIHR Global Health Research Group in Evidence to Policy pathway for Immunisation in China (EPICs; 16/137/109), the Key Program of the National Natural Science Foundation of China (82130093), the National Natural Science Foundation of China (92369118 and 82304202), and Shanghai Rising-Star Program (No. 22QA1402300).

## Author contributions

H. Y., J. Z., and M. A. designed the experiments. Y. L., J. Z., Q. Y., Q. W., X. Y., G. Z., K. D., Z. Z., N. L., X. Y., W. L., C. P., J. Z., and J. L. collected and cleaned the data. Y. L. and J. Z. analyzed the data. Y. L., J. Z., M. L., M. J., M. A., and H. Y. interpreted the results. Y. L., J. Z., and H. Y. wrote the manuscript. M. A. and M. J. edited the manuscript.

## Notes

### Author Declarations

All procedures performed in this study were approved by the ethics committees of the School of Public Health at Fudan University, Shanghai, China (Ref: 2020-11-0857-S2) and the London School of Hygiene and Tropical Medicine, London, UK (Ref: 22684-04), and were following the 1964 Helsinki declaration and its later amendments or comparable ethical standards. Informed consent was obtained from all individual participants included in the study (from a guardian if the participant was under 6 years old; from both a guardian and the study participant if the participant was 7-17 years old; from the participant if the participant was 18 years or above).

