## Supplementary materials for "Social contact patterns and their impact on the transmission of respiratory pathogens in rural China"

### Supplementary tables

#### Table S1 Representativeness of the samples.

| **Characteristics** | **Population of Anhua County, N (%) ^a^** | **Total sample size, N (%)** | **P-value ^b^** |
| --- | --- | --- | --- |
| **Overall** | 780,969 | 1,522 |  |
| **Sex** |  |  |  |
| Male | 399,177 (51.1) | 587 (38.6) | <0.001 |
| Female | 381,792 (48.9) | 935 (61.4) |  |
| **Age group** |  |  |  |
| 0-9 | 100,931 (12.9) | 411 (27.0) | <0.001 |
| 10-19 | 89,207 (11.4) | 235 (15.4) |  |
| 20-29 | 52,947 (6.8) | 167 (11.0) |  |
| 30-39 | 104,652 (13.4) | 181 (11.9) |  |
| 40-49 | 114,261 (14.6) | 206 (13.5) |  |
| 50-59 | 142,944 (18.3) | 141 (9.3) |  |
| 60-69 | 88,536 (11.3) | 100 (6.6) |  |
| 70+ | 87,491 (11.2) | 81 (5.3) |  |

^a^ 2020 Anhua census data.

^b^ Chi-square test

#### Table S2. Comparison between the mean number of contacts recorded on the day before the interview vs. another day (the nearest weekend/weekday)..

| **Variables^a^** | **aOR^b^** | **P-value^c^** |
| --- | --- | --- |
| **Diary date** |  |  |
| Yesterday | Ref |  |
| Another day | 1.11 (1.06, 1.16) | p<0.001 |
| **Education** |  |  |
| None | Ref |  |
| Primary school | 0.91 (0.78, 1.07) |  |
| Middle school | 0.86 (0.71, 1.04) |  |
| High school | 1.03 (0.82, 1.31) |  |
| College or above | 1.67 (1.28, 2.17) | p<0.001 |
| **Individual income (CNY)** |  |  |
| No income | Ref |  |
| <￥1 k | 1.01 (0.55, 1.87) |  |
| ￥1-9 k | 1.48 (1.17, 1.88) | p<0.001 |
| ￥10-29 k | 1.53 (1.28, 1.84) | p<0.001 |
| ≥￥30 k | 1.65 (1.37, 1.98) | p<0.001 |
| Not available | 2.42 (1.53, 3.83) | p<0.001 |
| Unwilling to answer | 1.24 (1.02, 1.5) | p<0.05 |
| Unknown/no response | 1.61 (1.05, 2.48) | p<0.05 |
| **Household size** |  |  |
| 1-3 | Ref |  |
| 4-6 | 1.16 (1.05, 1.28) | p<0.01 |
| ≥7 | 1.28 (1.1, 1.48) | p<0.01 |
| Unknown/no response | 0.63 (0.06, 6.65) |  |
| **Day type** |  |  |
| Workday | Ref |  |
| Weekend | 0.57 (0.55, 0.6) | p<0.001 |
| **Frequency of travels outside the village of residence** |  |  |
| Almost daily | Ref |  |
| At least once a week but not daily | 0.79 (0.69, 0.91) | p<0.001 |
| At least once a month but not each week | 0.73 (0.64, 0.84) | p<0.001 |
| Less than monthly, but not never | 0.71 (0.62, 0.82) | p<0.001 |
| Never | 0.7 (0.58, 0.86) | p<0.001 |
| Unknown/no response | 0.88 (0.7, 1.12) |  |
| Unwilling to answer |  |  |
| **Self-rated contact memory accuracy** |  |  |
| Very well | Very well | Ref |
| Well | Well | 1.19 (1.09, 1.29) |
| Moderate well | Moderate well | 1.25 (1.14, 1.37) |
| Not well | Not well | 1.18 (0.92, 1.51) |
| Unknown/no response | Unknown | 1.92 (1.38, 2.68) |

a Variables considered in the multivariate model except age, which was considered as smooth term in the model.

b Adjusted odds ratio.

c GAMM.

#### Table S3. Median number and interquartile range (IQR) of total, indoor, and outdoor contacts by participant’s characteristic.

| **Characteristics** | **Median (IQR)** | | |
| --- | --- | --- | --- |
|  | **Total** | **Indoor** | **Outdoor** |
| **Overall** | 7 (4, 14) | 6 (3, 10) | 0 (0, 1) |
| **Sex** |  |  |  |
| Male | 7 (4, 13) | 5 (3, 10) | 0 (0, 1) |
| Female | 7 (4, 14) | 6 (3, 11) | 0 (0, 1) |
| **Age group** |  |  |  |
| 0-2 yrs | 5 (4, 7) | 4.5 (3, 6) | 0 (0, 0) |
| 3-6 yrs | 6 (4, 11) | 6 (4, 10) | 0 (0, 0) |
| 7-19 yrs | 12 (6, 20) | 10 (5, 18) | 0 (0, 0) |
| 20-39 yrs | 8 (5, 14) | 6 (4, 10) | 0 (0, 2) |
| 40-59 yrs | 7 (4, 13) | 5 (3, 9) | 0 (0, 3) |
| 60-75 yrs | 5 (3, 8) | 4 (2, 6) | 0 (0, 1.8) |
| ≥76 yrs | 4 (2, 6) | 3 (2, 5) | 0 (0, 0) |
| **Education** |  |  |  |
| None | 6 (4, 10) | 5 (3, 8) | 0 (0, 1) |
| Primary school | 7 (4, 14) | 5 (3, 9) | 0 (0, 2) |
| Middle school | 10 (6, 17) | 9 (4, 14) | 0 (0, 1) |
| High school | 7 (4, 13.2) | 6 (3, 10) | 0 (0, 1) |
| College or above | 9.5 (5, 32) | 8 (4, 27.2) | 0 (0, 2) |
| **Occupation** |  |  |  |
| Enrolled in pre-kindergarten | 5 (4, 7) | 5 (3, 6) | 0 (0, 0) |
| Enrolled in kindergarten | 7 (4.8, 13) | 6 (4, 12) | 0 (0, 0) |
| Student | 12 (6, 20) | 10 (5, 17) | 0 (0, 0) |
| Self-employed | 15 (9, 24.8) | 12 (6, 21.5) | 0 (0, 1) |
| Farmer | 6 (3, 9) | 4 (2, 7) | 0 (0, 2) |
| School staff | 12 (5.2, 54.8) | 9.5 (5, 47) | 0 (0, 1.8) |
| Other staff | 15 (8.8, 34) | 10 (6, 15.5) | 0 (0, 4) |
| Others | 9 (5, 17) | 7 (4, 11) | 0 (0, 2) |
| Unemployed | 6 (3, 10) | 5 (3, 7) | 0 (0, 2) |
| **Individual income (CNY)** |  |  |  |
| No income | 6 (3, 10) | 5 (3, 7) | 0 (0, 2) |
| <￥1 k | 4.5 (3, 7.5) | 3 (2, 6) | 0 (0, 3) |
| ￥1-9 k | 7 (3, 15) | 4 (2, 8) | 0 (0, 4) |
| ￥10-29 k | 8 (5, 16) | 6 (3, 11) | 0 (0, 3) |
| ≥￥30 k | 9 (5, 22) | 8 (4, 16) | 0 (0, 2) |
| Not available | 8 (5, 16) | 7 (4, 13) | 0 (0, 0) |
| Unwilling to answer | 6 (3, 10) | 4 (2, 7) | 0 (0, 1.5) |
| Unknown/no response | 6 (4, 13) | 5 (4, 13) | 0 (0, 2) |
| **Household size** |  |  |  |
| 1-3 | 5 (3, 11) | 4 (2, 8) | 0 (0, 2) |
| 4-6 | 7 (5, 13) | 6 (4, 10) | 0 (0, 1) |
| ≥7 | 13 (7, 21) | 11 (6, 19) | 0 (0, 0) |
| Unknown/no response | 5 (5, 5) | 1 (1, 1) | 4 (4, 4) |
| **Number of years living in Anhua county** |  |  |  |
| <1 yrs | 6 (4, 9) | 5.5 (4, 9) | 0 (0, 0) |
| 1-5 yrs | 6 (4, 10) | 5 (4, 8) | 0 (0, 0) |
| 6-10 yrs | 8 (5, 15) | 7 (4, 12) | 0 (0, 0) |
| >10 yrs | 8 (4, 15) | 6 (3, 11) | 0 (0, 1) |
| Unknown/no response | 11 (5.5, 16.8) | 7.5 (3.8, 9.8) | 0.5 (0, 2) |
| **Day type** |  |  |  |
| Workday | 9 (5, 17) | 7 (4, 13) | 0 (0, 1) |
| Weekend | 6 (4, 11) | 5 (3, 8.2) | 0 (0, 1) |
| **Typical day** |  |  |  |
| Yes | 7 (4, 13) | 5 (3, 10) | 0 (0, 1) |
| No | 11 (7, 18) | 9 (6, 13) | 0 (0, 2) |
| Unknown/no response | 6 (3, 21) | 5 (0, 16) | 0 (0, 1) |
| **Weather** |  |  |  |
| Sunny | 7 (4, 13) | 5 (3, 10) | 0 (0, 1) |
| Cloudy | 7 (4, 15) | 6 (3, 12) | 0 (0, 1) |
| Rainy | 8 (4, 15) | 7 (3, 12) | 0 (0, 1) |
| Variable | 7 (5, 13) | 6 (3, 10) | 0 (0, 1) |
| Unknown/no response | 5 (3, 8.2) | 3 (2, 6.2) | 0 (0, 1) |
| **Frequency of travelling outside the village of residence** |  |  |  |
| Almost daily | 9 (5, 22) | 7 (4, 16) | 0 (0, 1) |
| At least once a week but not daily | 8 (5, 15.8) | 6 (4, 11) | 0 (0, 1) |
| At least once a month but not each week | 7 (5, 15) | 6 (3, 11) | 0 (0, 1) |
| Less than monthly, but not never | 7 (4, 11) | 5 (3, 9) | 0 (0, 1) |
| Never | 5 (3, 9) | 3 (2, 7) | 0 (0, 1) |
| Unknown/no response | 15 (9, 21.5) | 11 (7.8, 17.2) | 0 (0, 1) |
| **Underlying conditions** |  |  |  |
| Yes | 6 (3, 11) | 5 (2, 8) | 0 (0, 2) |
| No | 7 (4, 14) | 6 (3, 11) | 0 (0, 1) |
| Unknown/no response | 9.5 (6, 19.2) | 8.5 (5.2, 16.8) | 0 (0, 0) |
| Unwilling to answer | 6 (4, 12) | 6 (3, 11) | 0 (0, 1) |
| **Contact with animals** |  |  |  |
| Yes | 7 (4, 12) | 5 (3, 9) | 0 (0, 1) |
| No | 8 (4, 17) | 6 (3, 14) | 0 (0, 0) |
| Unknown/no response | 7.5 (5, 10) | 3.5 (1, 6) | 4 (4, 4) |
| **Self-reported accuracy of contact memory** |  |  |  |
| Very well | 6 (4, 10) | 5 (3, 8) | 0 (0, 1) |
| Well | 8 (4, 14) | 6 (3, 11) | 0 (0, 1) |
| Moderate well | 12.5 (7, 21.8) | 10 (4, 16) | 0 (0, 1) |
| Not well | 13 (6, 20) | 6 (4, 11) | 0 (0, 2) |
| Unknown/no response | 5 (3, 20) | 4 (0, 16) | 0 (0, 0) |
| **Health status** |  |  |  |
| Very well | 7 (4, 13) | 6 (3, 10) | 0 (0, 1) |
| Well | 8 (4, 16) | 6 (3, 12) | 0 (0, 1) |
| Moderate well | 8 (4, 15) | 6 (3, 11) | 0 (0, 2) |
| Not well | 5 (3, 12) | 5 (2, 9) | 0 (0, 1) |
| Unknown/no response | 6.5 (4, 16.2) | 4.5 (0.2, 14) | 0 (0, 0.8) |

### Supplementary figures

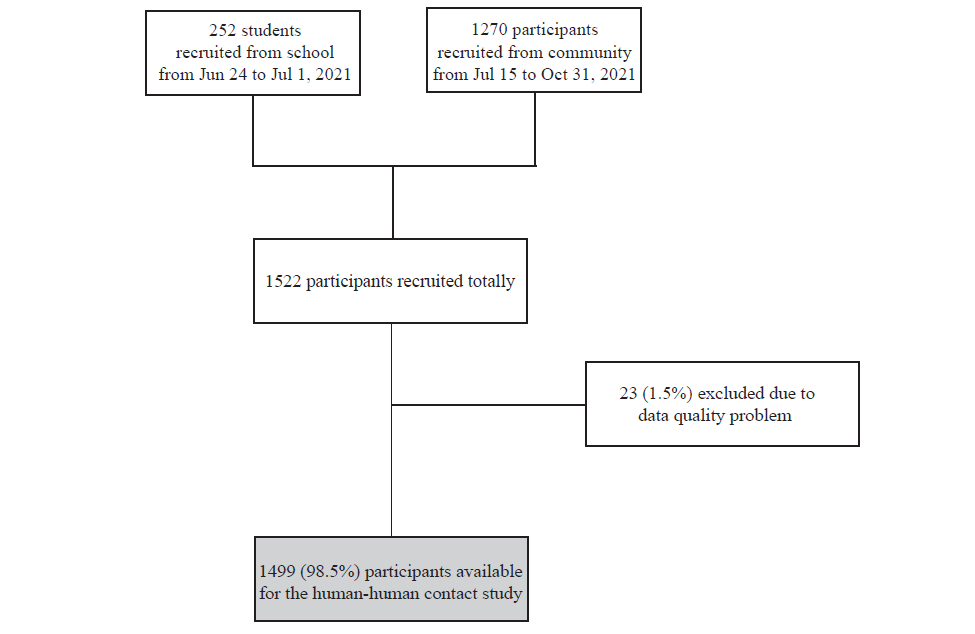

#### Figure S1 Flowchart of participants recruitment and determination of final sample.

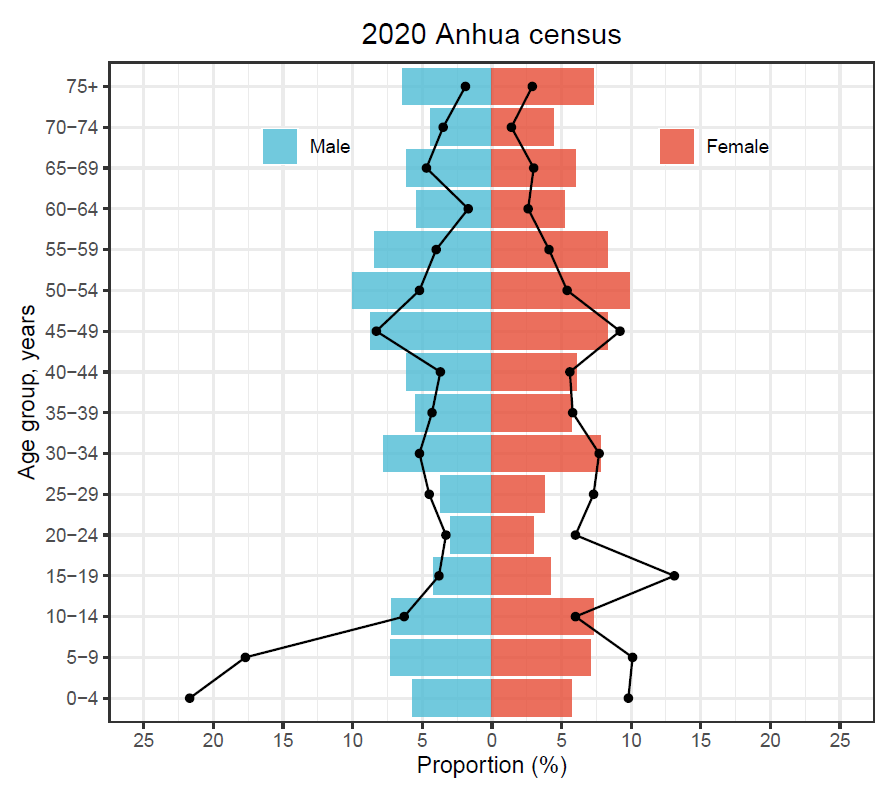

#### Figure S2 Representativeness of sample. Comparison between sample and 2020 census data by age and sex. Dots and lines denote the sample, bars denote the census data.

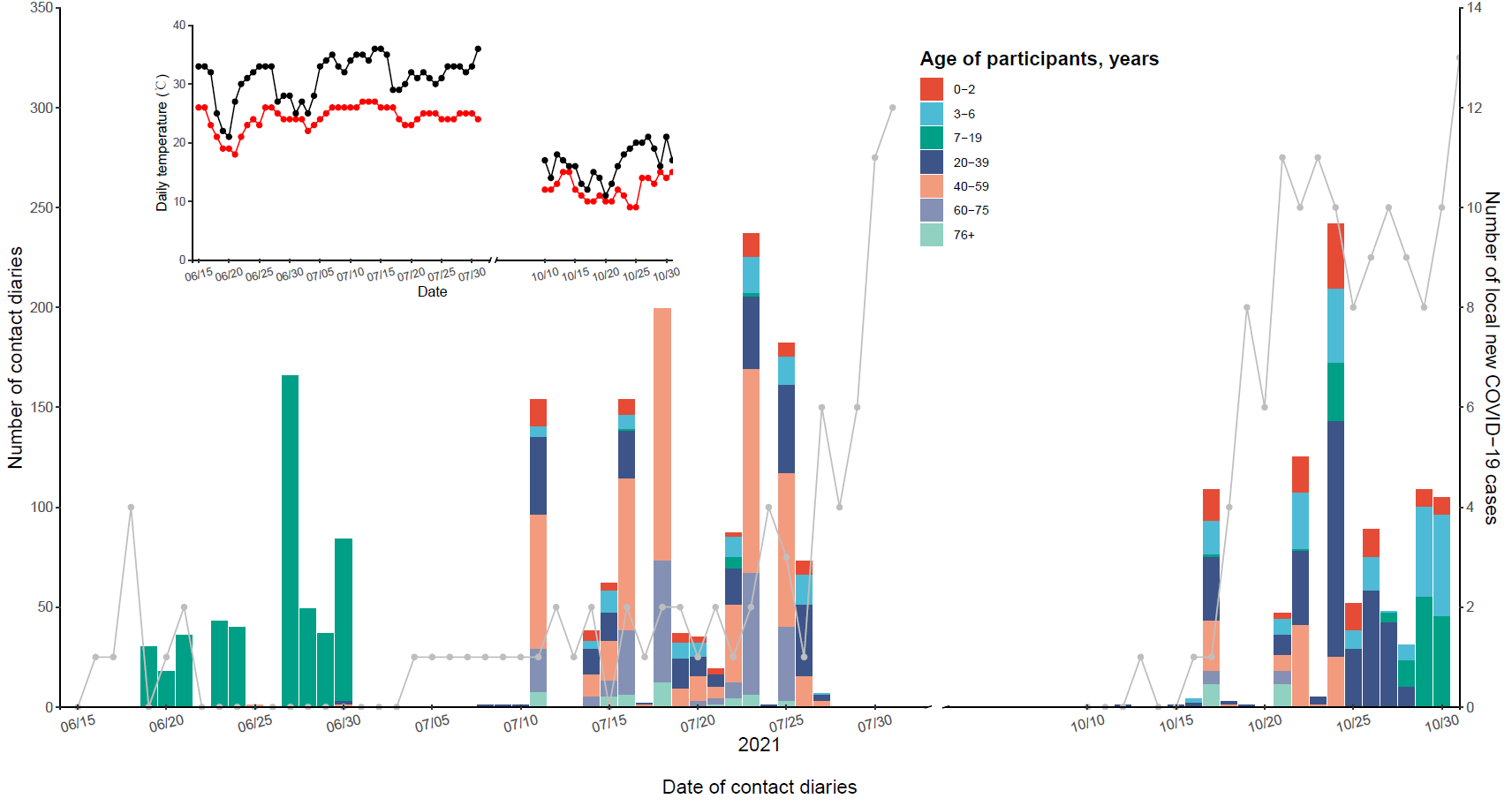

#### Figure S3 Number of surveys by date of social contact diary. The bars correspond to the number of diaries and the colors of bars represent age of participants (left vertical axis); the grey dots and lines represent the number of daily new local COVID-19 cases in Mainland China (right vertical axis). The inset shows the daily temperature during the survey; black dots and lines represent the daily maximum temperature, and the red dots and lines represent the daily minimum temperature.

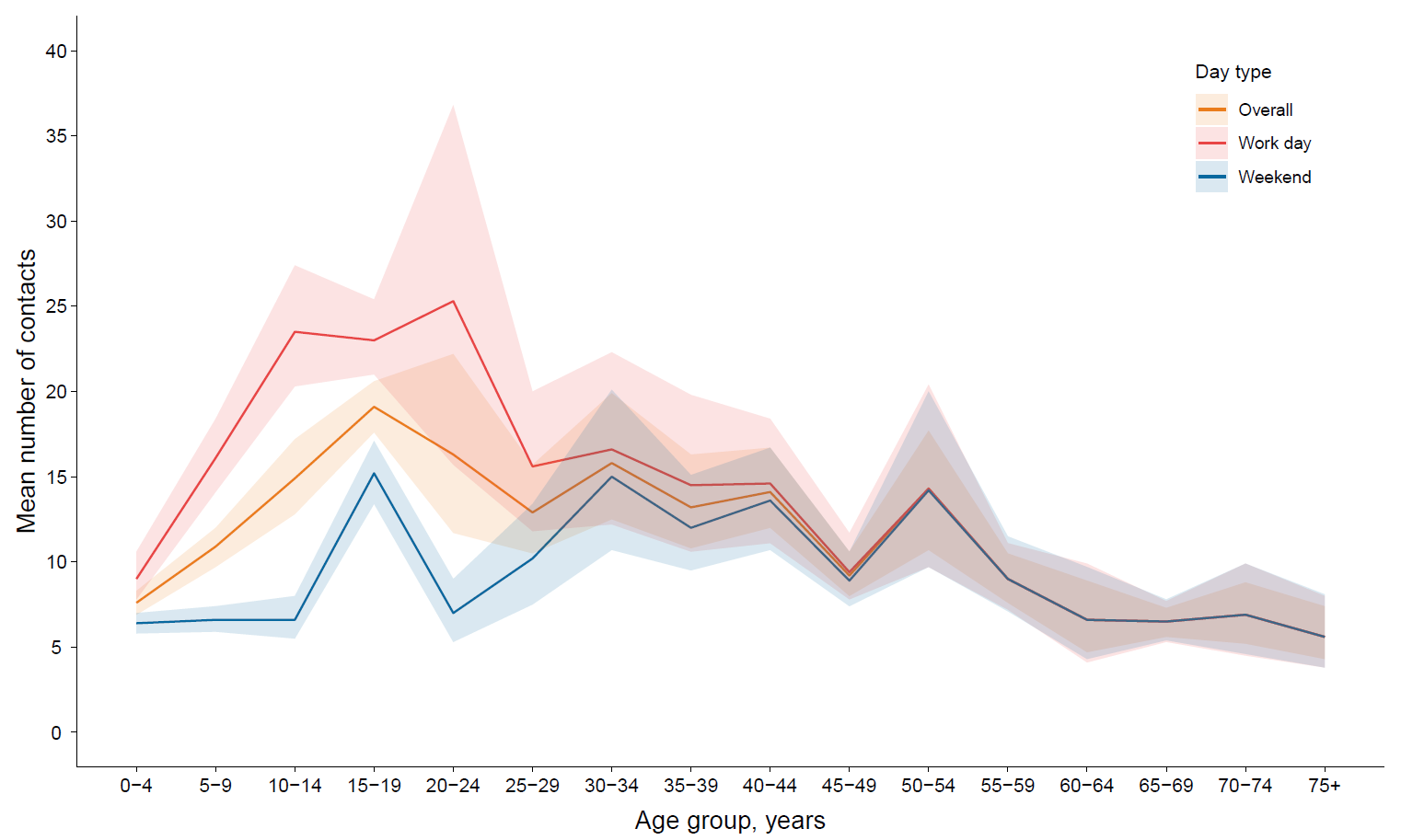

#### Figure S4 Mean number of contacts on workdays and weekends among participants of different ages. Shaded areas show 95%CI.

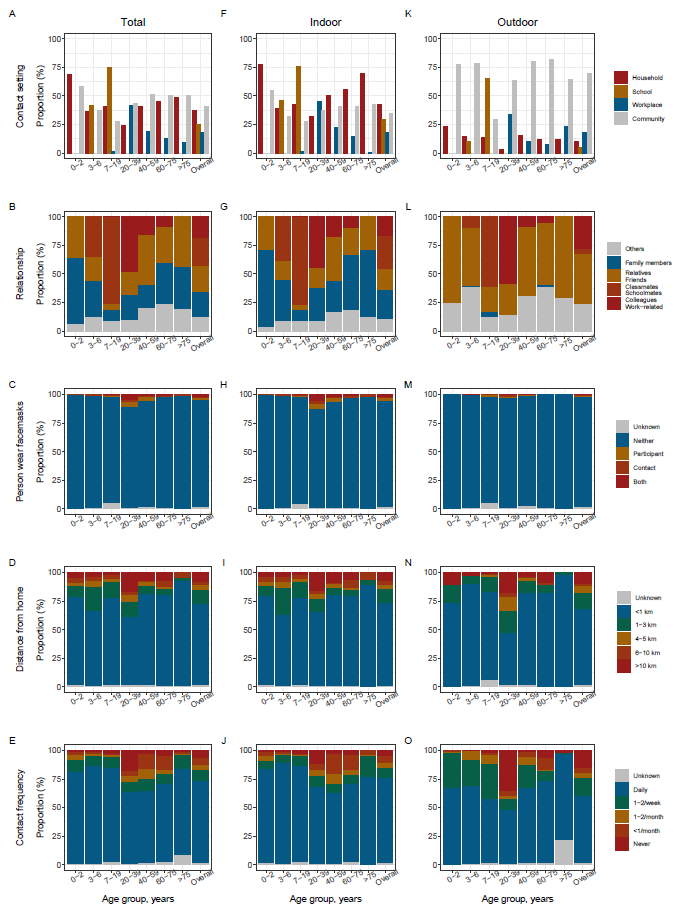

#### Figure S5 Details of total, indoor and outdoor contact number by age of participants.

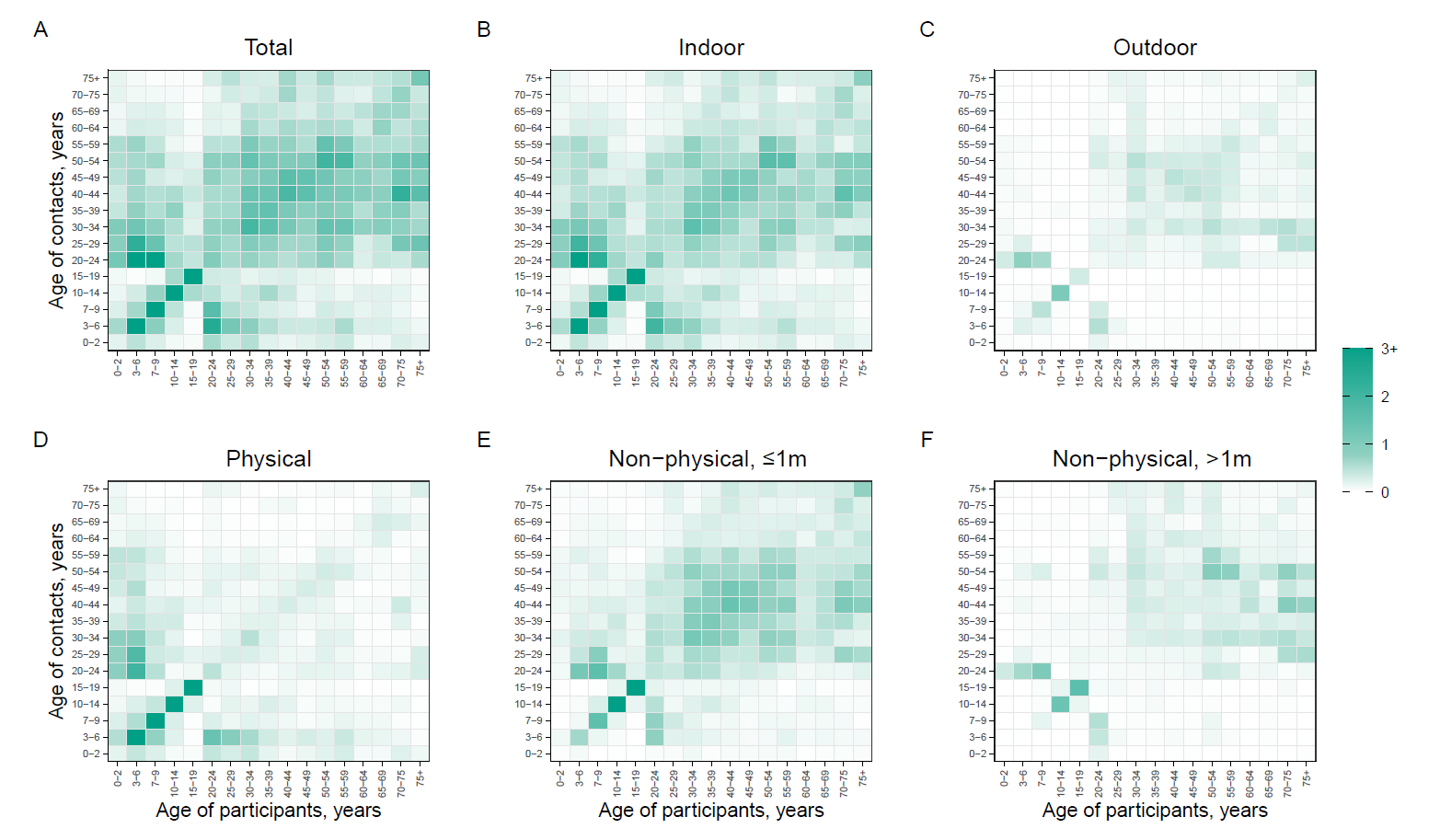

#### Figure S6 Original contact matrices.

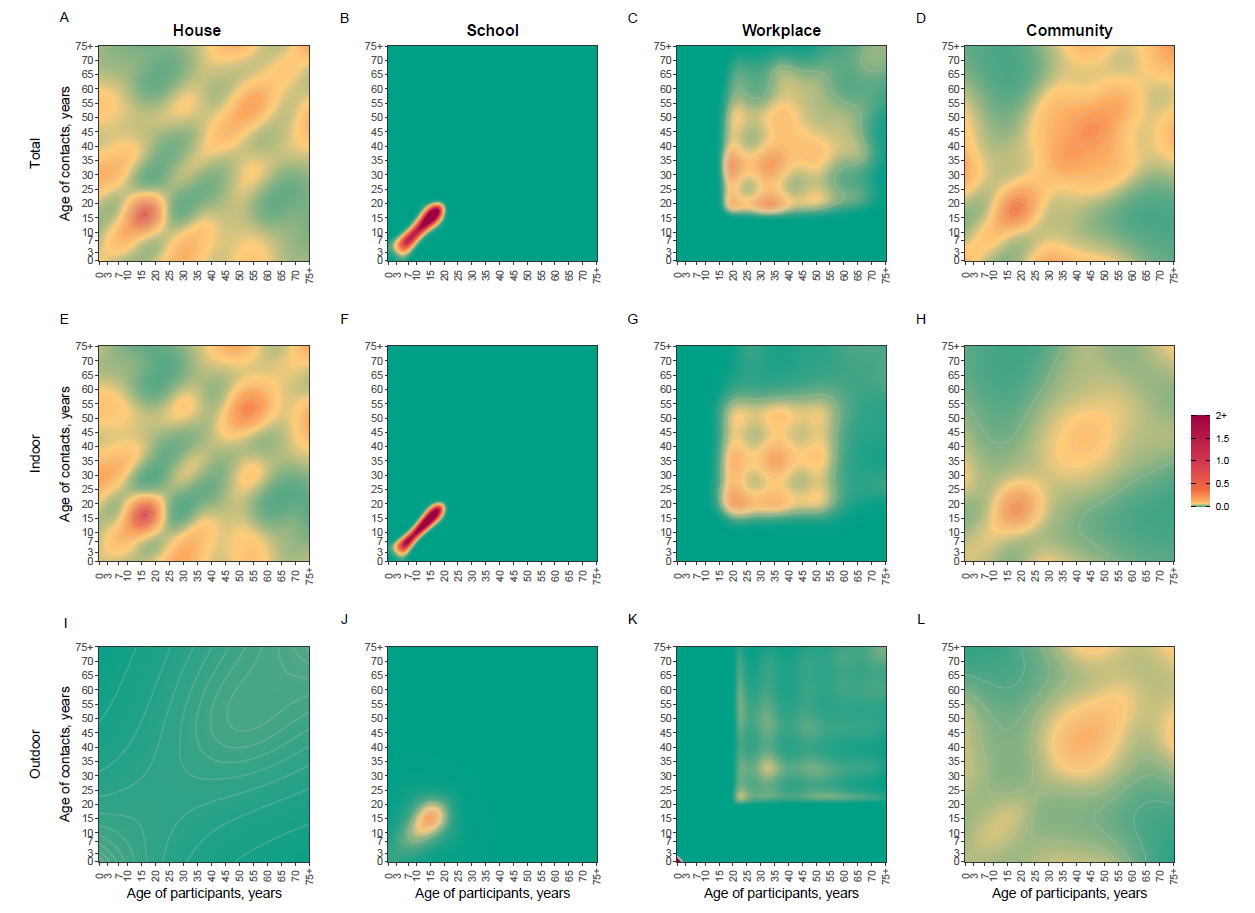

#### Figure S7 Contact matrices by settings.

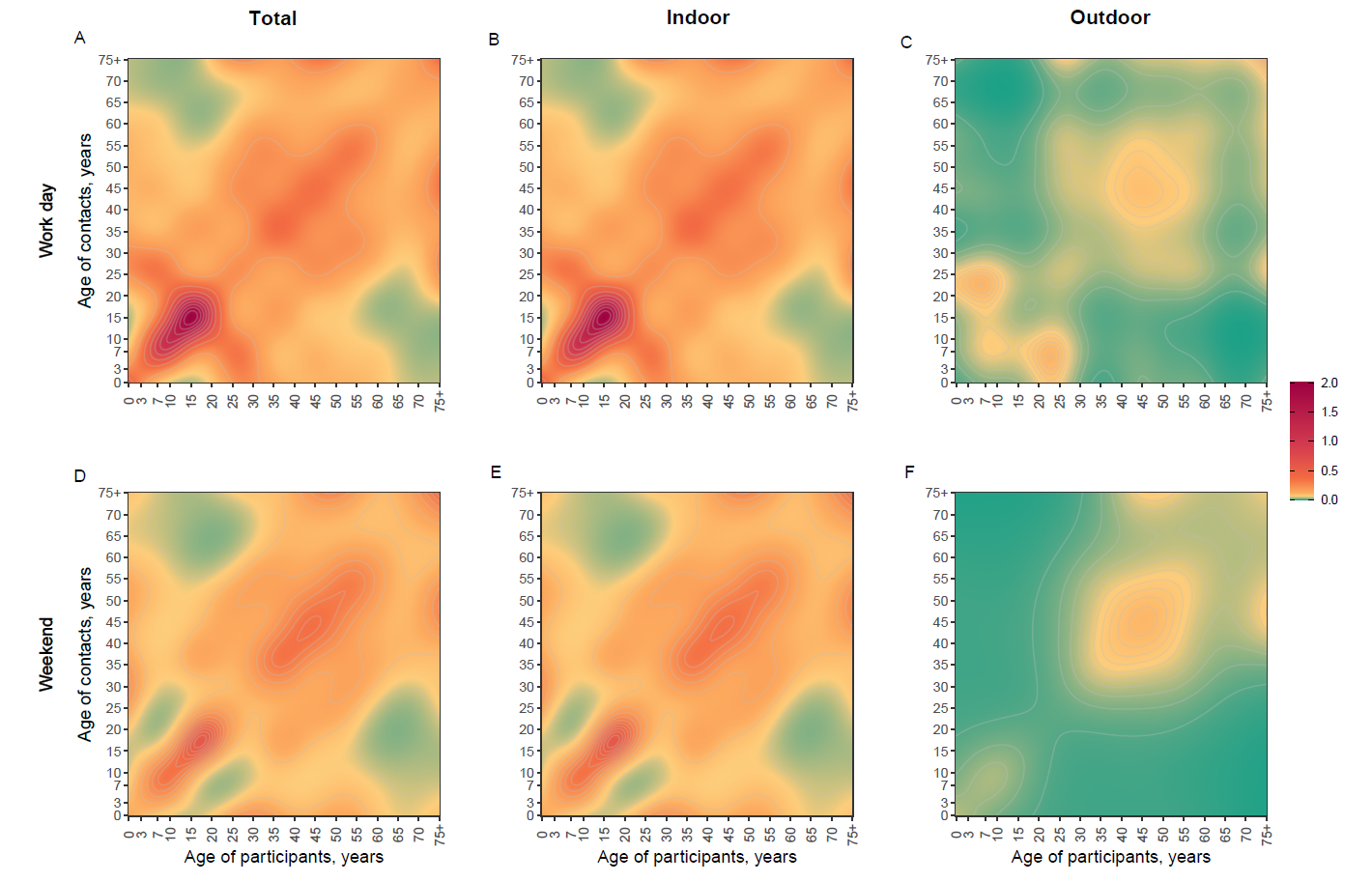

#### Figure S8 Contract matrices by day type.

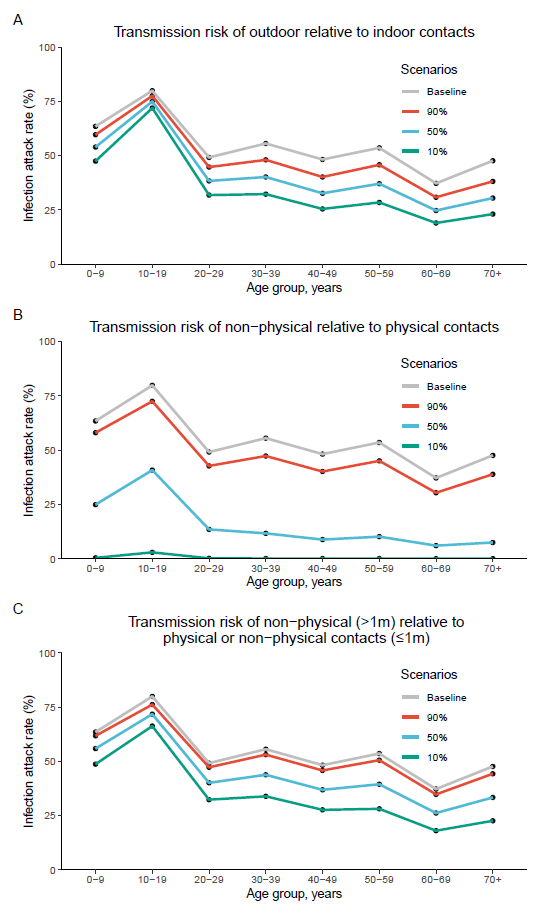

#### Figure S9 Estimated infection attack rate by age group with different modelled scenarios: (A) When varying the transmission risk of outdoor relative to indoor contacts. (B) When varying the transmission risk of non-physical relative to physical contacts. (C) When varying the transmission risk of non-physical contacts at more than 1-meter distance relative to physical or non-physical contacts at less than 1-meter distance.
